# Ongoing equipoise contributes to unchanged outcomes and significant morbidity following post-transplant cutaneous squamous cell carcinoma: A Multicentre Cohort Study

**DOI:** 10.1101/2025.06.02.25328493

**Authors:** THJ Crisp, E Peleva, CA Harwood, MJ Bottomley, the COAST Study Collaborative Group

## Abstract

**Introduction:** Cutaneous squamous cell carcinoma (CSCC) is the most common post-transplant malignancy, ultimately affecting up to half of all recipients. Historical cohort studies indicate that up to 25% of recipients develop further post-transplant CSCC within 12 months of their first. Post-transplant CSCC incurs doubled risk of metastasis and death compared to the general population and there is uncertainty regarding optimal secondary prevention strategies. Despite evolution of transplant cohort demographics and immunosuppressive regimens, there are no recent studies investigating outcomes after post-transplant CSCC.

**Methods:** A multi-centre retrospective cohort study was undertaken to evaluate current management practices and outcomes following first CSCC in kidney transplant recipients (KTR). Endpoints included development of further CSCC and adverse events including graft loss, solid-organ malignancy, metastatic CSCC, or death with a functioning graft. Multivariate survival modelling identified factors associated with immunosuppression reduction and increased risk of further CSCC and other adverse outcomes.

**Results:** 136 KTR from eight UK centres with first-ever CSCC diagnosed between 2016 and 2020 were identified. Median (IQR) follow-up was 39 (26-52) months, during which 48.5% developed further CSCC; these data were comparable to those from an international validation cohort. During follow up, 23.3% died, with malignancy being the most common cause of death. Post-CSCC management varied widely within and between centres, with 28.7% of KTR undergoing immunosuppression reduction.

**Conclusion:** Our findings demonstrate that outcomes after a first CSCC remain unchanged compared with historical studies, highlighting the urgent need for prospective interventional trials to establish the optimal secondary prevention strategies in this high-risk population.

**Lay Summary:** Cutaneous squamous cell carcinoma (CSCC), a type of skin cancer, is the most common cancer in organ transplant patients. Some patients develop more cancers or spread to other organs, but little is known about how best to manage transplant patients after CSCC to prevent further problems. To explore this, we studied a modern-day group of kidney transplant patients from eight UK hospitals after their first CSCC, reviewing how they were treated and what happened afterwards. We found that up to half developed another CSCC within three years. There was no consistent treatment between hospitals, showing that there is uncertainty about the best way to care for these patients. Many patients died or had other serious health problems. These results are similar to older studies from the past 20 years, suggesting little improvement in outcomes. We conclude that transplant patients with their first CSCC remain at high risk of further cancers and other complications. More research is urgently needed to reduce these risks and improve care.

**Visual Abstract:** 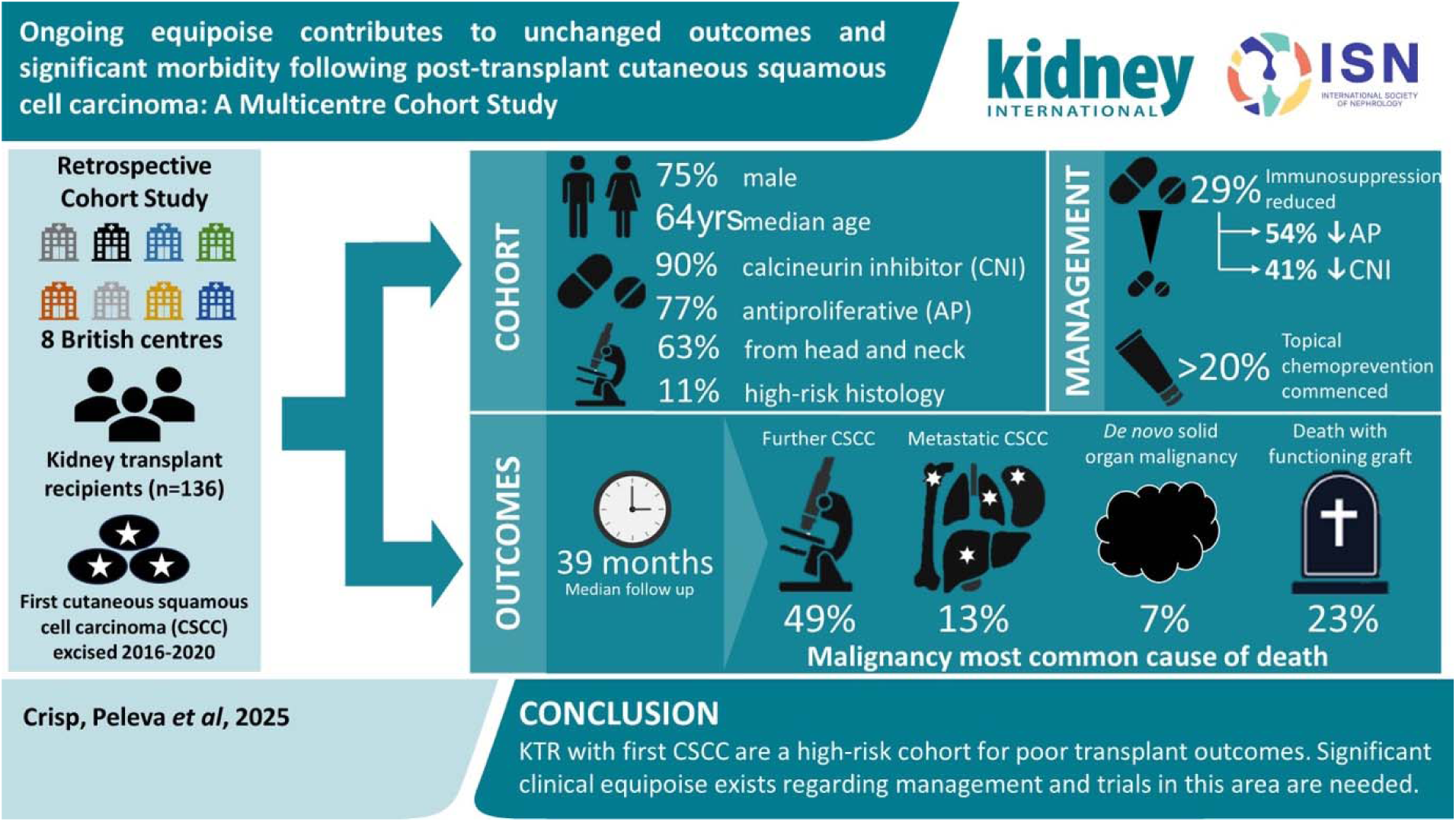

## Introduction

While short-term outcomes after kidney transplant have improved,^1^ long-term outcomes remain suboptimal. Half of kidney transplant recipients (KTR) will die with a functioning graft,^2^ with cancer remaining a leading cause of mortality.^3^ Skin cancer, 80% of which is cutaneous squamous cell carcinoma (CSCC), is the most common post-transplant malignancy, eventually affecting up to half of all solid organ transplant recipients (SOTR).^4,5^ Risk factors for post-transplant CSCC include older age, Fitzpatrick skin type I-II, increasing duration of immunosuppression and cumulative ultraviolet radiation exposure.^6^ Historical studies suggest that a quarter of SOTR with a first (primary) CSCC develop another within a year, rising to 60% within five years.^7^ Serial post-transplant CSCC are associated with increased risk of metastasis and solid organ malignancy (SOM).^5,8,9,10^

Despite the burden of post-transplant CSCC, clinical equipoise remains regarding strategies to limit further disease (secondary prevention).^11^ This uncertainty is driven by a lack of data from prospective interventional studies and limitations of existing observational cohorts, including their single centre nature, long study periods subject to era effects, the pooling of different SOTR types, and reliance on registry data lacking sufficient granularity. The lack of risk stratification tools identifying KTR at greatest risk of further disease for targeted use of secondary prevention approaches may also contribute.^11^

No recent studies have evaluated management or outcomes after CSCC in SOTR, obscuring contemporary practices regarding secondary prevention. Over the last twenty years, the mean age of KTR at transplant has increased by over a decade.^12^ In parallel, immunosuppressive protocols have evolved, with widespread adoption of antibody induction therapy, reduced corticosteroid use, and replacement of azathioprine and ciclosporin with mycophenolate mofetil (MMF) and tacrolimus, respectively. Modern immunosuppressive regimens may reduce the risk of primary CSCC,^13,14^ although this may be offset by an ageing transplant population and increased CSCC baseline risk, evidenced by conflicting epidemiological trends regarding primary CSCC incidence in SOTR.^15–17^ The effect of these demographic and management shifts on outcomes after CSCC remains unknown.

Extrapolation from interventional studies evaluating mammalian target of rapamycin inhibitors (mTORi) for secondary CSCC prevention suggests immunosuppression modulation may be most efficacious when implemented after the first-ever CSCC.^4,18^ It is unknown whether immunosuppression modification after first CSCC is widely used in real-world practice, especially as mTORi use may be limited by patient intolerance.^4,18,19^ There is a similar paucity of real-world data regarding the use of systemic and topical chemoprevention after first CSCC.

We hypothesise that lack of evidence has led to widespread variation in secondary prevention approaches after CSCC,^11^ undertaking the Contemporary Outcomes After Skin cancer in Transplant (COAST) study to assess both modern-day management and outcomes after first CSCC in KTR.

## Methods

COAST was a multicentre, retrospective cohort study identifying patients from eight renal centres in the south of England and Wales, including five transplanting centres. Centres are anonymised for reporting. Consecutive adults with a functioning kidney (or kidney and pancreas) transplant at time of their first-ever CSCC, diagnosed between 1^st^ January 2016 and 31^st^ December 2020, were included. We excluded patients with prior diagnosis of CSCC, or history of other organ or stem-cell transplant. Follow-up began on the date of the first CSCC histology report and finished at the last transplant or dermatology clinic appointment (if lost to follow-up), death, or 31^st^ December 2022 (whichever occurred first). Patients were additionally censored in the event of graft loss, defined as the initiation of dialysis.

Demographic, histopathological and clinical data were collected from local electronic patient records using a standardised data collection proforma (Supplementary Table S2). Data were cross-checked by a nephrologist and a dermatologist prior to anonymisation and compilation by the co-ordinating centre. CSCC were retrospectively staged using the Brigham and Women’s Hospital (BWH) classification, the 8^th^ edition of the American Joint Committee on Cancer (AJCC8)/International Union Against Cancer (UICC8) staging system, the Actinic Damage and Skin Cancer Index (AD-SCI) and the British Association of Dermatologists’ SCC guidelines (BAD).^20–23^ Multiple staging criteria were included due to limited data on their performance in immunosuppressed cohorts and uncertainty regarding the optimal criteria. Where multiple index lesions were excised, the highest risk lesion (by BWH staging) was used for survival analyses.

Early management was defined as any documented change in immunosuppression within six months of CSCC diagnosis. Immunosuppression intensity was characterised as decreased (‘immunosuppression reduction’, ISR) if dose reduction or cessation of an agent was undertaken. A switch of agent was considered as unchanged immunosuppression intensity. Initiation of topical treatments for pre-malignant actinic keratoses with potential chemopreventative activity (5-fluorouracil/imiquimod), destructive therapies (cryotherapy/curettage/cautery), and systemic chemoprevention (nicotinamide/acitretin) within this timeframe were also recorded, based on clinic letters and/or prescription records.

An international cohort was assembled to validate the COAST study findings regarding outcomes following a first CSCC. All adult SOTR with a first-ever CSCC diagnosed at the Brigham and Women’s Hospital, USA, between 1st January 2012 and 31st December 2020, were included. Patients were excluded if they had history of CSCC or stem cell transplant. Patients were also excluded if the index lesion was an CSCC-in situ without evidence of invasion, basosquamous carcinoma, or non-cutaneous SCC, or if they had missing data. The follow-up period was calculated as the date of the first CSCC histology report to the last skin cancer-related clinic, death, or 9^th^ April 2025 (whichever occurred first).

### Statistical analysis

Descriptive statistics were used to summarise clinical and histopathological characteristics. Continuous variables were reported using median and interquartile range (IQR) for non-parametric data and mean and standard deviation for parametric data. Baseline characteristics and management strategies across the eight study centres were compared by Pearson’s chi-squared test with a Monte Carlo simulation. Association between baseline characteristics and early ISR was assessed using the Mann–Whitney U test for continuous variables and Fisher’s exact test for categorical variables. Statistically significant factors were evaluated further using multivariate logistic regression modelling.

Survival and time-to-event data were analysed using the ‘survival’ package in R and visualised by Kaplan-Meier (KM) univariate modelling. KM curves were used to estimate the cumulative incidences of further CSCC and poor outcomes. Influence of clinico-pathological variables on patient outcomes was assessed using Cox proportional hazards models, constructed based on pre-specified clinical hypotheses and findings from exploratory univariate analyses. Study outcomes were ‘further CSCC’ development, and a composite ‘all poor outcomes’ including graft loss, further CSCC, SOM, metastatic CSCC, or death with a functioning graft (DWFG). Further CSCC was defined as a new primary CSCC; excluding recurrences within existing excision scars. In the case of further CSCC, multivariate modelling was conducted with competing risk analysis, applying the Fine-Gray method for sub-distribution proportional hazards modelling implemented via the ‘cmprsk’ R package. Death and graft loss were treated as competing risks.^24^ Results are reported as hazard ratios (HR) with 95% confidence intervals (CI). As a sensitivity analysis, survival models for the composite poor outcome were repeated after excluding further CSCC as an event (‘poor outcomes without CSCC’).

Statistical significance was set at 0.05 (two-sided). Statistical analysis was performed using R version 4.3.2 via RStudio, using the most recent versions of the appropriate packages.^25^ All plots were created using the ‘ggplot2’ package.

This report was written in accordance with STROBE reporting guidelines (Supplementary Table S3).^26^

### Ethical approval

The COAST study was conducted in accordance with the Declaration of Helsinki and was approved by the NHS Human Research Authority (Ethics reference:22/HRA/3782). Participant consent was not required as the study used routinely collected clinical information only.

## Results

Initial data were provided for 153 patients. 17 were excluded after review (Supplementary Table S4) leaving 136 eligible patients for downstream analysis (Figure 1a).

**Figure 1.**
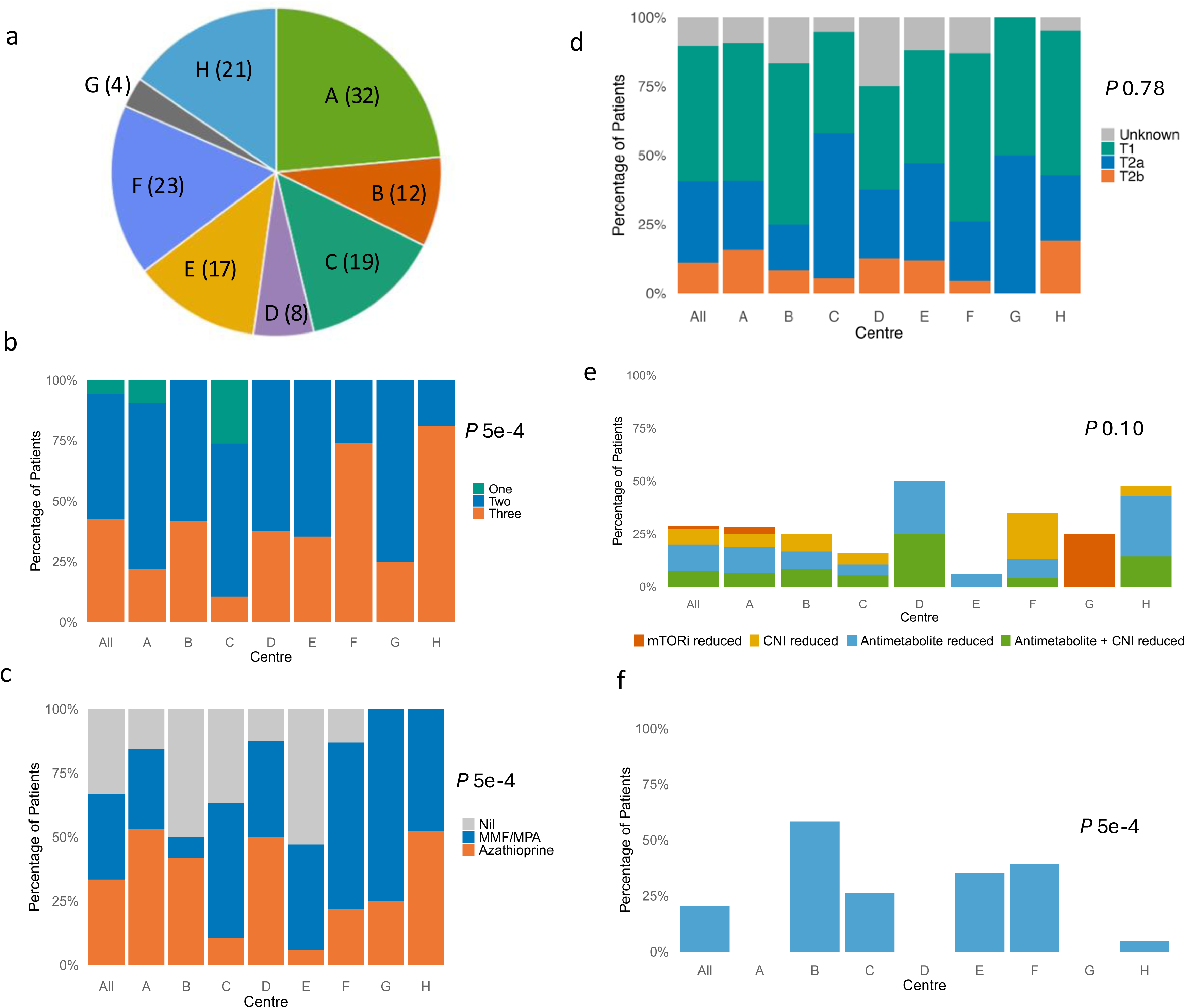

### Baseline characteristics

Table 1 summarises cohort features at time of first CSCC. Notably, most patients were male (75.0%), white (97.7%) and had a median (IQR) age of 64 (57-72) years. The median eGFR was 41 (27-55) ml/min/1.73m^2^. 15.7% had previously been treated for biopsy-proven acute rejection.

**Table 1.**
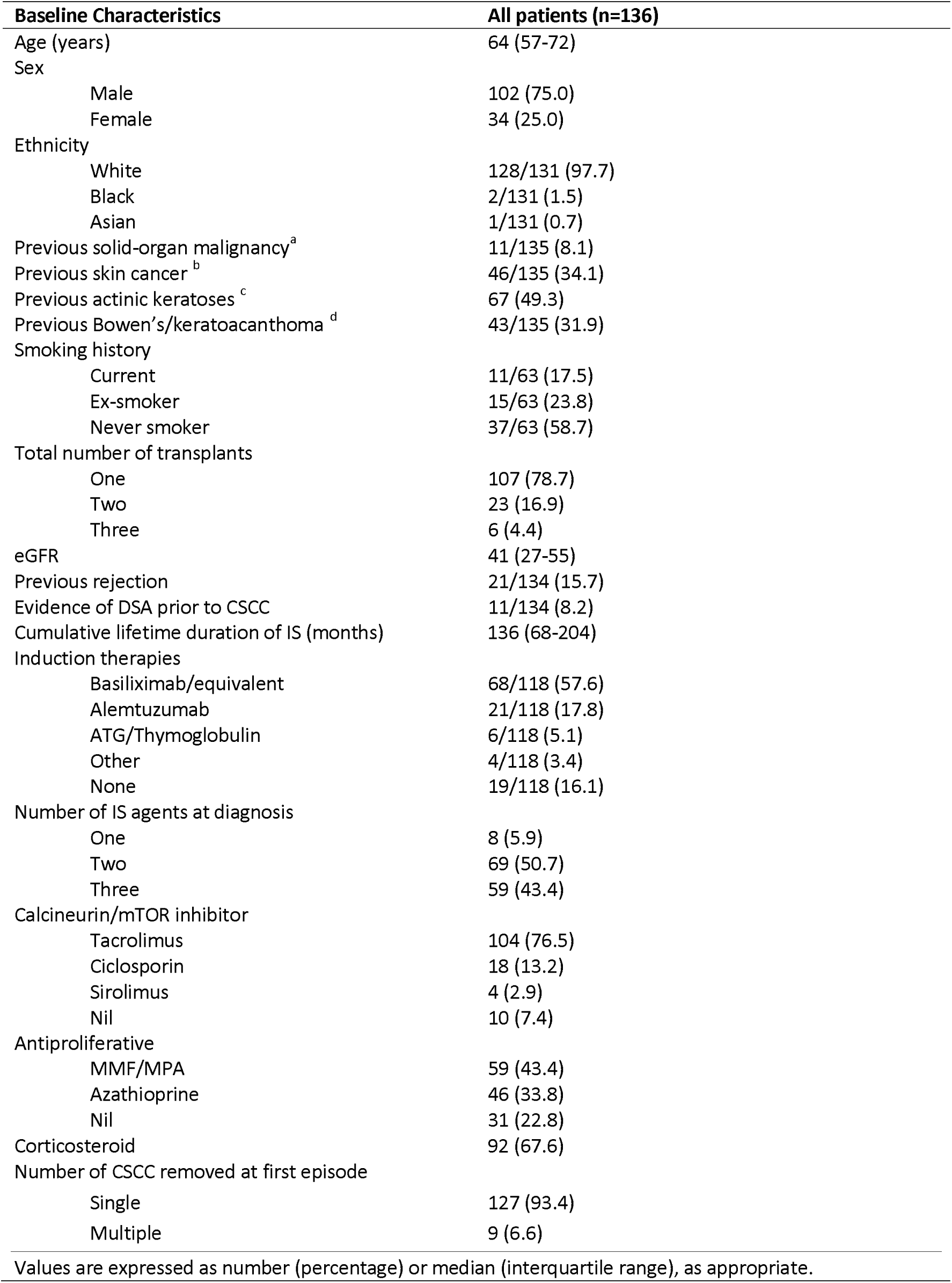
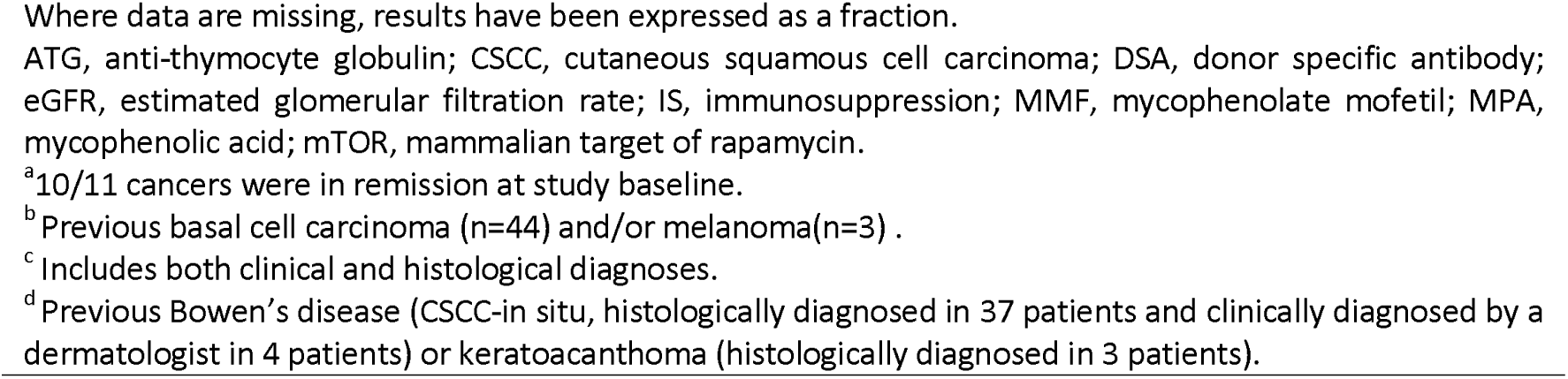
Baseline characteristics of 136 patients diagnosed with first cutaneous squamous cell carcinoma (CSCC)

| Baseline Characteristics | All patients (n=136) |
| --- | --- |
| Age (years) | 64 (57-72) |
| Sex |  |
| Male | 102 (75.0) |
| Female | 34 (25.0) |
| Ethnicity |  |
| White | 128/131 (97.7) |
| Black | 2/131 (1.5) |
| Asian | 1/131 (0.7) |
| Previous solid-organ malignancy<sup>a</sup> | 11/135 (8.1) |
| Previous skin cancer<sup>b</sup> | 46/135 (34.1) |
| Previous actinic keratoses<sup>c</sup> | 67 (49.3) |
| Previous Bowen's/keratoacanthoma<sup>d</sup> | 43/135 (31.9) |
| Smoking history |  |
| Current | 11/63 (17.5) |
| Ex-smoker | 15/63 (23.8) |
| Never smoker | 37/63 (58.7) |
| Total number of transplants |  |
| One | 107 (78.7) |
| Two | 23 (16.9) |
| Three | 6 (4.4) |
| eGFR | 41 (27-55) |
| Previous rejection | 21/134 (15.7) |
| Evidence of DSA prior to CSCC | 11/134 (8.2) |
| Cumulative lifetime duration of IS (months) | 136 (68-204) |
| Induction therapies |  |
| Basiliximab/equivalent | 68/118 (57.6) |
| Alemtuzumab | 21/118 (17.8) |
| ATG/Thymoglobulin | 6/118 (5.1) |
| Other | 4/118 (3.4) |
| None | 19/118 (16.1) |
| Number of IS agents at diagnosis |  |
| One | 8 (5.9) |
| Two | 69 (50.7) |
| Three | 59 (43.4) |
| Calcineurin/mTOR inhibitor |  |
| Tacrolimus | 104 (76.5) |
| Ciclosporin | 18 (13.2) |
| Sirolimus | 4 (2.9) |
| Nil | 10 (7.4) |
| Antiproliferative |  |
| MMF/MPA | 59 (43.4) |
| Azathioprine | 46 (33.8) |
| Nil | 31 (22.8) |
| Corticosteroid | 92 (67.6) |
| Number of CSCC removed at first episode |  |
| Single | 127 (93.4) |
| Multiple | 9 (6.6) |

Values are expressed as number (percentage) or median (interquartile range), as appropriate.
Where data are missing, results have been expressed as a fraction.
ATG, anti-thymocyte globulin; CSCC, cutaneous squamous cell carcinoma; DSA, donor specific antibody; eGFR, estimated glomerular filtration rate; IS, immunosuppression; MMF, mycophenolate mofetil; MPA, mycophenolic acid; mTOR, mammalian target of rapamycin.
<sup>a</sup>10/11 cancers were in remission at study baseline.
<sup>b</sup>Previous basal cell carcinoma (n=44) and/or melanoma(n=3) .
<sup>c</sup>Includes both clinical and histological diagnoses.
<sup>d</sup>Previous Bowen's disease (CSCC-in situ, histologically diagnosed in 37 patients and clinically diagnosed by a dermatologist in 4 patients) or keratoacanthoma (histologically diagnosed in 3 patients).

A third of the cohort (34.1%) had a previous non-CSCC skin cancer diagnosis (predominantly basal cell carcinoma, BCC) while 11 patients (8.1%) had a history of SOM. Over half (60.0%) had a previously documented clinical or histological diagnosis of a pre-malignant skin lesion including actinic keratosis, Bowen’s disease (CSCC-in situ) or keratoacanthoma. Comparing baseline characteristics across centres, there were significant differences (P <0.05, chi-squared test) in the proportions of patients with previous rejection episodes, immunosuppression regimens (induction therapy, antiproliferative, steroid use, and number of agents), year of first CSCC (2016-2020), and whether multiple CSCC were removed. Sex, smoking status, and history of BCC were not significantly different between centres (Supplementary Table S5).

### Immunosuppression at first CSCC

The median cumulative duration of immunosuppression at first CSCC was over 11 years (136 months, IQR 68-204). Over 80% of patients, where this information was available, had previously received induction therapy, with the majority receiving CD25-targetting monoclonal antibodies. Patients were most commonly on two or three immunosuppressive agents at CSCC diagnosis. Most KTR (89.7%) were taking a calcineurin inhibitor (CNI), most commonly tacrolimus (76.5%). The majority (77.2%) were taking an antiproliferative agent, most commonly a mycophenolic acid-based preparation (43.4%). Overall, only one-third of patients were receiving azathioprine. Two-thirds were receiving oral corticosteroids at first CSCC. Immunosuppression regimens varied significantly across centres (Figure 1b-c, Supplementary Table S5), in terms of number of agents, antiproliferative and corticosteroid use (P 5e-4, chi-squared test).

### Histological characteristics of first CSCC

First CSCC histopathological characteristics are detailed in Table 2. Nine KTR (6.6%) had multiple CSCC excised at their index episode. 147 tumours in total were excised at index presentation, most frequently located on the head and neck (63.3%) and upper limb (25.2%). Four-fifths of tumours had clear excision margins, rising to 92.8% following re-excision. There was a discrepancy in risk staging depending on the system used: 14 tumours were high-risk by both BWH (defined as ≥T2b) and AJCC8 (≥T3); 17 tumours were high-grade by AJCC8 only (of which 11 were T2a and six were T1 by BWH); one was high grade by BWH only (AJCC8 T2). The proportion of patients with a first ‘high-grade’ CSCC varied across centres (AJCC8 range 5-50% and BWH 0-20%, Figure 1d), although this did not reach statistical significance (P> 0.05, chi-squared test).

**Table 2:** Characteristics of first cutaneous squamous cell carcinoma (CSCC)

| Characteristics | Number of tumours (n=147) |
| --- | --- |
| Location |  |
| Head and neck | 93 (63.3) |
| Trunk | 12 (8.2) |
| Upper limb | 37 (25.2) |
| Lower limb | 5 (3.4) |
| Diameter (mm) | 12 (9-20) |
| Depth (mm) | 3 (1.9-5) |
| Invasion beyond the subcutis | 8/128 (6.3) |
| Differentiation |  |
| Well | 40/145 (27.6) |
| Moderate | 78/145 (53.8) |
| Poor | 27/145 (18.6) |
| PNI | 8/130 (6.2) |
| LVI | 3/141 (2.1) |
| Clear margins after excision | 117/146 (80.1) |
| Clear margins after re-excision | 128/139 (92.8) |
| BWH |  |
| T1 | 77/133 (57.9) |
| T2a | 41/133 (30.8) |
| T2b | 15/133 (11.3) |
| AJCC8/UICC8 |  |
| T1 | 91/134 (67.9) |
| T2 | 11/134 (8.2) |
| T3 | 32/134 (23.9) |
| AD-SCI |  |
| 4 | 85/134 (63.4) |
| 5 | 16/134 (11.9) |
| 6 | 33/134 (24.6) |
| BAD |  |
| High risk | 95 (70.4) |
| Very high risk | 40 (29.6) |

Values are expressed as number (percentage) or median (interquartile range), as appropriate.
Where data is missing, results have been expressed as a fraction.
AD-SCI, Actinic Damage and Skin Cancer Index; AJCC8/UICC8, the 8th edition of the American Joint Committee on Cancer/International Union Against Cancer staging systems; BAD: British Association of Dermatologists' SCC guidelines; BWH, Brigham and Women's Hospital classification; LVI, lympho-vascular invasion; PNI, perineural invasion

### Early Management after CSCC

ISR was undertaken within six months of first CSCC in 39 patients (28.7%), with reduction of antiproliferative, CNI or mTORi in 17 (43.6%), 10 (25.6%) and 2 (5.1%) patients, respectively. Additionally, 10 patients (25.6%) had reduction of both antiproliferative and CNI. Of the 27 patients who had reduction of an antiproliferative agent, the majority were receiving azathioprine (20 patients, 74.1%), with the remainder on MMF (25.9%). One patient was converted to an mTORi. CSCC was specifically cited as the reason for ISR in 21 patients (53.8%). Other cited reasons included undesirable trough levels, recurrent infections, and adverse effects.

ISR frequency varied significantly among patients on different drug regimens: ISR was observed in 21/46 (45.7%) of patients receiving azathioprine, 13/59 (22.0%) receiving mycophenolate-based regimens, 35/122 (28.9%) receiving CNI, and 3/4 (75%) receiving mTORi (P 0.01, Fisher’s test).

There was no statistically significant difference in the proportions of patients who underwent early ISR across study centres (P 0.10, chi-squared test); however, the pattern of ISR varied within and between centres (Figure 1e). A centre’s transplanting status did not significantly affect frequency or type of ISR (P 0.36 and *P* 0.06, Fisher’s test, respectively). Further evaluating factors associated with ISR (Table 3), patients with a high-grade first CSCC by BWH criteria (≥T2b) were more likely to have ISR (OR 4.6, *P* 0.01, Fisher’s test). Conversely, those with a previous BCC were less likely to have ISR (OR 0.17, *P* 0.0005, Fisher’s test). There was a trend towards a lower eGFR in patients undergoing ISR (33 vs 41ml/min/1.73m^2^, *P* 0.09, Mann-Whitney U test).

**Table 3.**
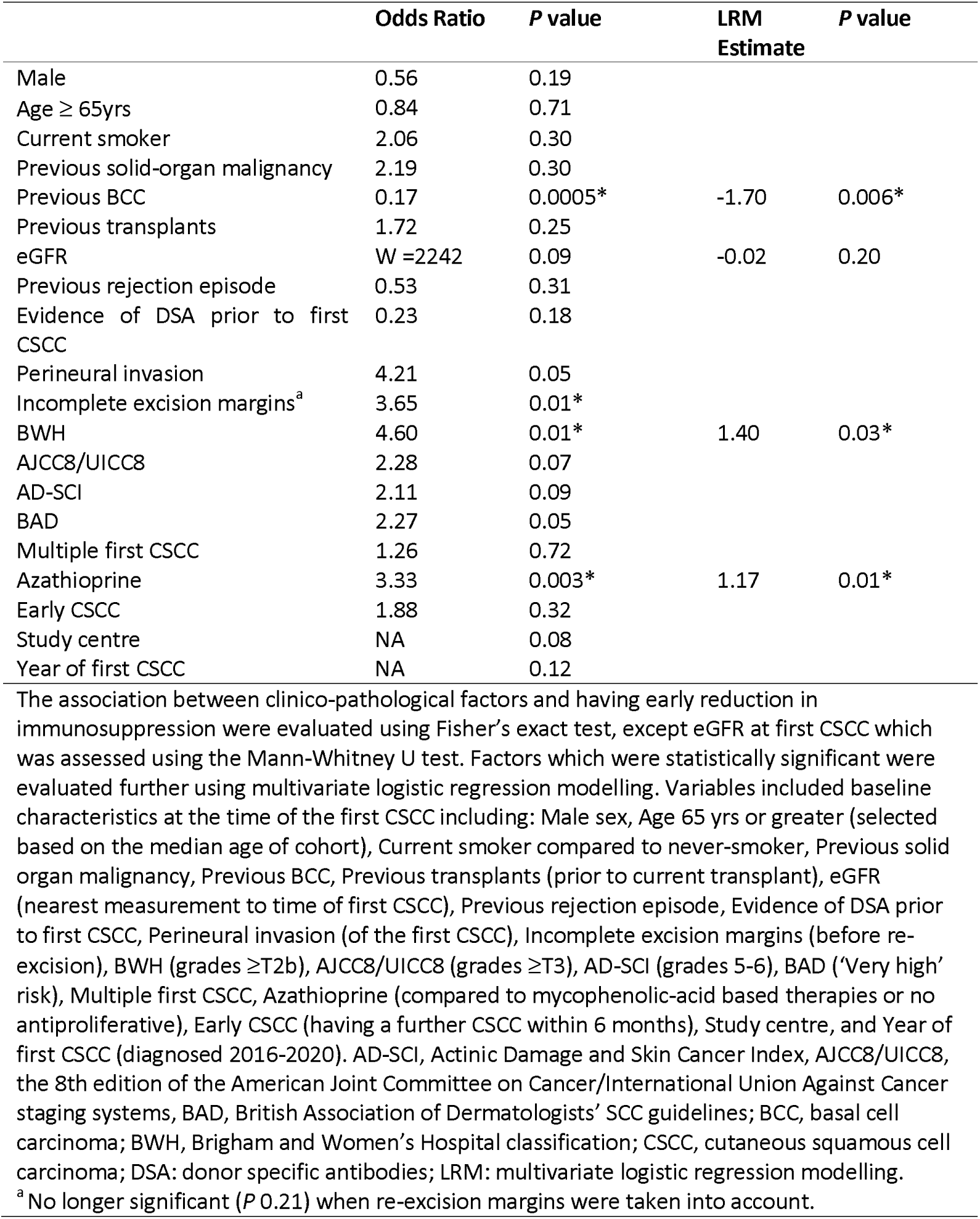
Factors associated with early reduction in immunosuppression.

28 patients (20.6%) had documented initiation of topical therapies within six months of first CSCC, of which all but one started 5-fluorouracil cream (5-FU). The frequency of initiation varied significantly across centres (Figure 1f, *P* 5e-4, chi-squared test). Six patients (4.4%) started systemic chemoprevention.

### Outcomes

Study outcomes are shown in Table 4 and summarised in Figure 2a-b. During a median (IQR) follow-up of 39 (26-52) months, 66 patients (48.5%) developed further CSCC, with tumour burden ranging from 1-13 (median 2) further lesions. The estimated cumulative incidence (95% CI) of further CSCC was 24.6% (16.8%-31.6%), 47.8% (37.7%-56.3%), and 65.2% (45.9%-77.6%) at one, three, and five years, respectively (Figure 2c), with a median (IQR) interval between first and second CSCC of 13 (8-27) months. This cumulative incidence is comparable to historical studies (Figure 2d).^7,10,27^

**Figure 2.**
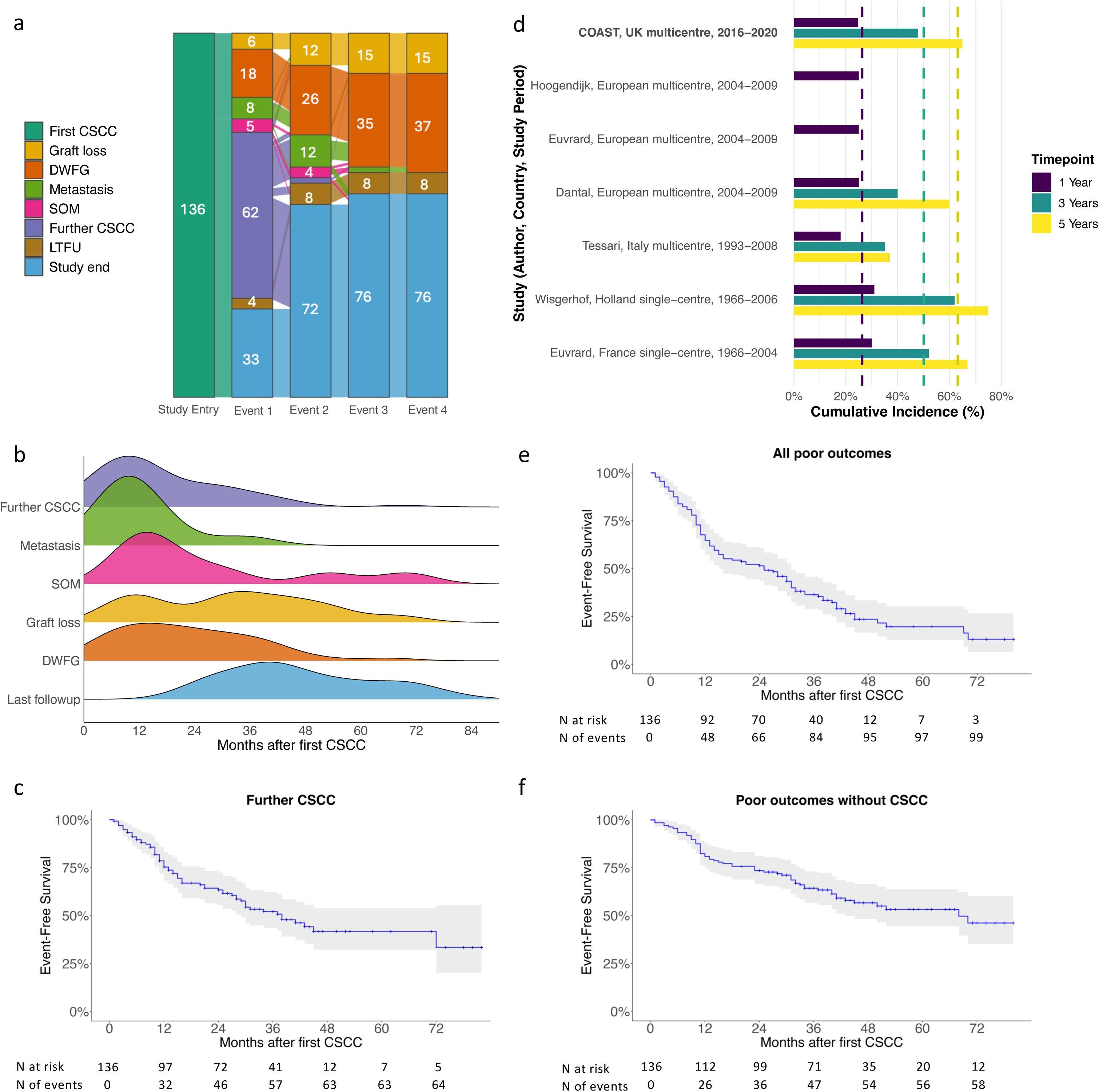

**Table 4.** Outcomes during follow-up.

| Outcome | All patients (n=136) |
| --- | --- |
| Follow-up (months) | 39 (26-52) |
| All poor outcomes <sup>a</sup> | 99 (72.8) |
| Death | 38 (23.3) |
| Time to death (months) | 20 (11-32) |
| Causes of death |  |
| CSCC-related | 11 (28.9) |
| Infection | 6 (15.8) |
| CVD | 6 (15.8) |
| Other malignancy | 2 (5.3) |
| Decompensated liver disease | 1 (2.6) |
| Other | 2 (5.3) |
| Unknown | 10 (26.3) |
| Graft loss | 17 (12.5) |
| Time to graft loss (months) | 34 (18-45) |
| Biopsy-proven rejection | 1/132 (0.8) |
| New DSA | 3/110 (2.7) |
| Further CSCC | 66 (48.5) |
| Median time to further CSCC (months) | 13 (8-27) |
| Metastatic CSCC | 18/135 (13.3) |
| Time to metastatic CSCC (months) | 11 (7-12) |
| Solid-organ malignancy <sup>b</sup> | 10 (7.4) |
| Time to solid-organ malignancy (months) | 16 (14-27) |

Values are expressed as number (percentage) or median (interquartile range), as appropriate.
CSCC, cutaneous squamous cell carcinoma; CVD, cardiovascular disease; DSA, donor-specific antibody.
<sup>a</sup> Composite outcome including graft loss, new CSCC, solid-organ malignancy, metastasis or death with a functioning graft.
<sup>b</sup> Solid organ malignancies included gastro-intestinal malignancy (n = 3), cancer of transplanted kidney (n=2), post-transplant lymphoproliferative disorder (n=2), breast cancer (n=1), tonsillar carcinoma (n=1), pleomorphic dermal sarcoma (n=1), and pseudomyxoma peritonei (n=1).

18 patients (13.3%) developed metastatic CSCC, with 15 cases confirmed on histopathology and three diagnosed on imaging. Eleven had metastases limited to the parotid gland and lymph nodes, while the remaining seven had distant spread. One-third of CSCC metastases (n=6) occurred in patients with a solitary prior CSCC. Median (IQR) interval to metastasis diagnosis was 11 (7-12) months, with 89% of patients developing metastases within two years of their first CSCC. The estimated median survival for patients with metastatic CSCC was 23 months (95% CI: 17 months to not estimable), and their estimated one-, three-, and five-year survival was 83.0%, 39.1% and 32.6%, respectively.

38 patients (23.3%) died during follow-up, representing the most common cause of premature censoring. The median (IQR) time from first CSCC to DWFG was 20 (12-32) months. The estimated all-cause mortality rates in the cohort were 8.1%, 24.0%, and 31.4% at one, three, and five years, respectively. The most common cause of DWFG was malignancy-related (n= 13, 34.2%), of which 11 (84.6%) were directly attributed to CSCC, yielding a crude CSCC-specific mortality rate of 8% (8.7% excluding unknown causes of death). Other major causes of DWFG included infection (15.8%) and cardiovascular disease (15.8%).

Graft loss occurred in 17 patients (12.5%), with a median (IQR) interval of 34 (18-45) months after first CSCC. ISR was not associated with an increased risk of graft loss during follow-up (*P* 0.23, Fisher’s test). The only episode of biopsy-proven rejection during follow-up occurred in a patient who had not undergone ISR.

*De novo* SOM was diagnosed in ten patients (7.4%) during follow-up, with a median (IQR) interval from first CSCC of 16 (14-27) months. 70% of cancers occurred within two years of the first CSCC, and half of those with new SOM had previously had multiple CSCC excised. Six of these patients died during follow-up. The estimated one-, three-, and five-year survival for those diagnosed with SOM was 100%, 66.7%, and 50%, respectively. Of those patients with new SOM, four (40%) progressed to metastatic disease. One patient with history of breast cancer preceding the first CSCC also developed metastases at 14 months, and this was presumed to be breast cancer recurrence. The estimated one-, three-, and five-year survival for those diagnosed with metastatic cancer (secondary to CSCC or SOM) during follow-up were 86.7%, 36.5%, and 26.1%, respectively.

99 patients (72.8%) developed at least one composite poor outcome (graft loss, further CSCC, SOM, metastatic CSCC or DWFG) during follow-up (Figure 2a), with most (66.7%) poor outcomes occurring within two years of the first CSCC (Figure 2b). 58 patients (42.9%) developed a poor outcome when further CSCC was removed from the composite (Figure 2f).

### Risk factors for further lesion development

We evaluated characteristics present at time of first CSCC diagnosis associated with the development of further CSCC and poor outcomes, which might assist in risk stratification (Figure 3).

**Figure 3.**
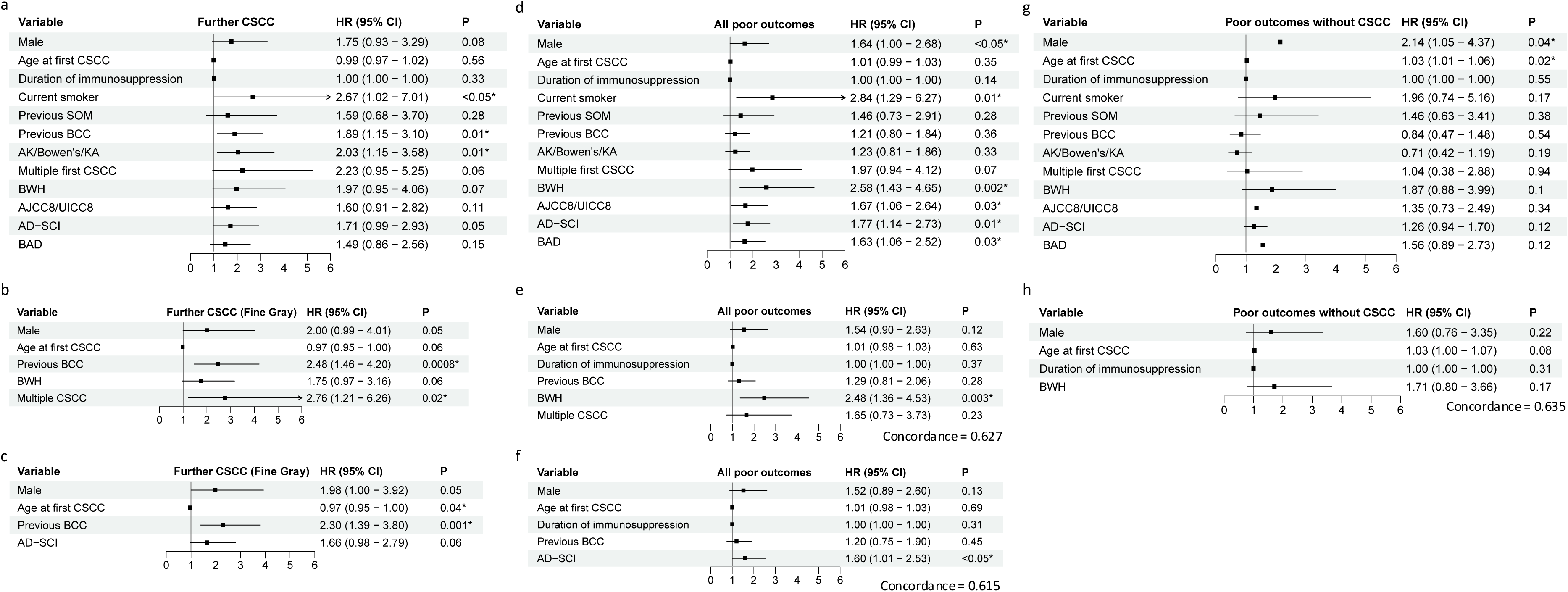

Smoking (hazard ratio, HR 2.7), previous BCC (HR 1.9), and pre-malignant skin lesions (HR 2.0) were predictive for developing further CSCC on KM univariate analysis (Figure 3a). Male sex, high-risk CSCC, by either BWH (HR 2.0) or AD-SCI (HR 1.7), and multiple first CSCC (HR 2.2) showed a trend towards increased risk, without reaching significance (P 0.06-0.08).

As smoking status was unknown for 53.7% of our cohort and documented history of pre-malignant lesions was likely underreported, both were excluded from further analysis. For multivariate modelling, we included statistically significant variables, or those trending towards significance (P <0.10), as well as age and duration of immunosuppression, which are important clinical risk factors.^6^ History of BCC and multiple first CSCC were independent predictors for further CSCC on multivariate analysis, accounting for competing risks (Figure 3b-c).

Male sex (HR 1.6), smoking (HR 2.8) and high-risk CSCC (AJCC8/UICC8 HR 1.7, BWH HR 2.6, AD-SCI HR 1.8, BAD HR 1.6) were significant predictors for ‘all poor outcomes’ (Figure 3d). Multiple first CSCC trended towards significance (HR 2.0, *P* 0.07). On multivariate analysis, high-risk CSCC (by BWH or AD-SCI) remained an independent predictor for ‘all poor outcomes’ (Figure 3e-f). As the most frequent poor outcome was further CSCC, analyses were repeated excluding this. Only male sex (HR 2.1) and age at first CSCC (HR 1.03) remained significant predictors of poor outcomes after excluding further CSCC, and there was a trend for high-risk CSCC by BWH (HR 1.9, Figure 3g).

### International validation of outcomes

To confirm our findings regarding significant morbidity after first post-transplant CSCC, we undertook validation in an external cohort of 79 North American SOTR (including 46 KTR) with a first CSCC during 2012-2020. Demographics and CSCC characteristics were similar between the cohorts (Supplementary Tables S6 and S7). The validation cohort had a lower cumulative duration of immunosuppression (64 vs 136 months), but a greater proportion were receiving triple-immunotherapy (57.5% vs 42.6%). Notably, index CSCC were more frequently low-risk (90.6% by BWH and 94.1% by AJCC). Follow-up duration was longer in the validation cohort (median 59 months, IQR 38-93). Comparably poor outcomes were observed after the first CSCC (Supplementary Table S8). The cumulative incidence of further CSCC at one, three, and five years were 29.4%, 58.3% and 72.1%, respectively (Supplementary Figure S1a). These were not significantly different from our cohort (P >0.05). Analysis of outcomes using KM curves showed no significant differences between KTR and other SOTR (Supplementary Figure S1).

## Discussion

Malignancy is a leading cause of post-transplant morbidity and mortality, with CSCC specifically a major contributor to this. However, little is known about outcomes after first CSCC in KTR in the modern era of immunosuppression. Additionally, uncertainty regarding optimal management has been highlighted by the lack of consensus from a recent Delphi panel on managing low-risk CSCC in SOTR or on immunosuppression modification after first CSCC.^21^

This retrospective study is the first to evaluate real-world management and outcomes after the first CSCC in a contemporary cohort of KTR, and the largest ever study conducted over a single era of practice. We identify that this cohort develops a significant burden of morbidity after their index lesion, with up to half developing further CSCC and the majority developing one or more of the composite ‘all poor outcomes’ within five years (many within two years). DWFG was observed in around a quarter of patients, with malignancy being the leading cause, surpassing both infection and cardiovascular mortality combined. Metastatic CSCC occurred in 13.3%, typically within two years of the first lesion, with poor survival. Significant variation in early management approaches was noted from both dermatology and transplant practitioners. Whilst optimal secondary prevention strategies for this high-risk cohort remain unclear, we have identified common risk factors for both further CSCC and poor outcomes, namely male sex, multiple/high-grade CSCC and history of BCC, which may identify patients most likely to benefit from targeted intervention.

The multicentre design of this study overcomes limitations from prior studies by identifying a cohort over a relatively short period. This approach permits the evaluation of management practices without confounding from era effect or local practice variations, often present in single-centre studies. Validation in an external cohort confirmed equally poor outcomes internationally, independent of the healthcare setting.

Three similar studies were undertaken in North European SOTR in the early 2000s, and provide a historical comparator.^7,10,27^ Contemporary KTR are approximately a decade older at time of their first CSCC, reflecting the increasing age of patients at transplant.^12^ Male-predominance in the cohort was unchanged, reflecting sex-specific differences in chronic kidney disease prevalence alongside greater environmental CSCC risk factor exposure amongst men.^28^ The interval between transplant and first CSCC is broadly unchanged at around 11 years. In the historical studies, the majority of patients were receiving three-agent immunosuppression, with three-quarters receiving azathioprine. Almost no KTR received antibody induction therapy.^7,10,27^ In contrast, induction therapy was used in the majority of our cohort. Observational studies suggest T-cell depleting induction therapy, received by a minority of our cohort, may specifically be associated with increased post-transplant skin cancer risk.^29^ Azathioprine, associated with enhanced CSCC risk in preclinical and observational studies,^30^ was only used by one-third of patients at the time of first CSCC reflecting a shift towards mycophenolic acid-based antiproliferative therapy since the 2000s. Importantly, this suggests that strategies and guidelines aimed at lowering CSCC risk through reduced azathioprine use or transitioning to alternative agents are less likely to apply to KTR in the future.

Up to half of KTR developed another CSCC within three years, representing a cumulative incidence almost identical to previous studies.^7,10,27^ Age is a major risk factor for CSCC development regardless of immunosuppression status, and the increasing age of KTR may be hypothesised to increase CSCC burden. Conversely, newer immunosuppressive regimens are associated with lower carcinogenic risk.^13,14,17^ We conclude that, despite changes in KTR demographics and immunosuppressive regimens over the last twenty years, secondary outcomes following CSCC remain effectively unchanged.

One strength of the COAST study was the granular nature of patient-level data collected, permitting detailed evaluation of other relevant transplant outcomes. A composite poor outcome measure was used to capture the frequency of adverse events and highlight the significant morbidity and mortality among KTR after first CSCC. These outcomes were identified as important and combined into a composite outcome because all are associated with a deleterious impact upon graft or patient survival. During the study period, 7.4% of patients developed a SOM, with a median interval of 16 months from first CSCC to diagnosis, and one-third died within three years. For those who developed metastatic CSCC, the three-year survival was 39.1%, substantially lower than the 60% three-year metastatic CSCC-specific survival reported previously,^31^ which may reflect the increased age and/or comorbidity-burden of contemporary KTR.^12^ Nearly one-quarter of our cohort (23.3%) died during the study period, with a median time to DWFG of 20 months. By comparison, a prospective cohort study of CSCC in the general population, which included immunosuppressed patients and SOTR, reported a median overall survival of 51.8 months.^32^ Our study included graft loss as a key outcome, recognising its significance to both patients and physicians.^33^ Fear of graft loss has been cited as a reason for hesitance to reduce immunosuppression following CSCC; reassuringly, our data showed no increase in biopsy-proven rejection or graft loss among those who underwent ISR.

Our study demonstrates significant intra- and inter-centre variation in immunosuppression modulation and chemoprevention use after a first CSCC. Centres appeared to have different strategies for secondary prevention, including ISR (range 6%-50%), topical (range 0%-58%) and systemic chemoprevention (range 0%-25%). The use of topical therapies is likely underreported in clinical records, particularly among patients attending specialised dermatology clinics, where regular self-directed 5-FU treatment may not have been systematically documented. Our data provides the first real-world evidence for the lack of consensus around management after a first CSCC, as indicated in a recent Delphi study.^21^

Difficulty stratifying KTR at highest risk of poor outcomes after first CSCC may represent a significant cause of ‘therapeutic momentum’ (defined as the reluctance to reduce therapy when continuation is not needed or not supported by evidence)^34^ regarding immunosuppression modulation for many clinicians. To help address this, we identified readily-available clinico-pathological factors at the time of first CSCC which are predictive of poor outcomes. Factors predictive of further CSCC likely relate to cumulative UV exposure, whilst high-risk histology may be a read-out of shared genetic and immunological host factors that predispose to further malignancy and/or mortality. These features may be clinically utilisable to target KTR who may benefit most from prompt intervention.

The broad range of management strategies reflects the ongoing lack of a consensus and limited evidence base for practice in this area, and likely contributes significantly to the apparent lack of improvement in outcomes seen in the contemporary era. Notably, only one patient was converted to an mTORi after diagnosis of first CSCC, despite randomised trial data indicating greatest efficacy when this intervention is deployed at this stage.^4,18^ Low uptake of this approach likely reflects real-world hesitancy, resulting from the high burden of adverse effects reported in clinical studies, leading to discontinuation in up to 80%.^19^

Limitations of our study include its retrospective nature, which introduces inherent biases such as the potential for unmeasured confounding and reliance on the completeness of medical records. No comparison cohort was available to evaluate outcomes of matched KTR without a first CSCC. Our international cohort showed similarly poor outcomes but had limited numbers of KTR.

We conclude that KTR with a first CSCC are at high risk for poor outcomes, leading to significant mortality and morbidity, and this remains unchanged from historical studies despite advances in transplant medicine. Continued clinical equipoise regarding secondary prevention management in this population has led to a range of different strategies and the potential for undertreatment. We identify factors that may permit a stratification approach to highlight KTR who may benefit most from secondary prevention strategies after their first CSCC, whilst also urgently advocating for prospective studies to establish optimal management for this high-risk cohort.

## Supporting information

Supplemental Table S2 - Data collection proforma

## Data Availability

Data availability statement
MJB is the dataset custodian. Data will be made available, upon reasonable request, by contact with the corresponding author.

## Acknowledgements

The authors would like to thank all members of the research team at participating centres who contributed to record collection. MJB is a career development fellow and is supported by the Chinese Academy of Medical Sciences (CAMS) Innovation Fund for Medical Science (CIFMS), China (grant number: 2018-I2M-2-002). EP is supported by the Wellcome Trust HARP PhD Studentship (grant number 318534/Z/24/Z). Earlier versions of these results were presented as abstracts at the British Association of Dermatologists’ annual meeting (2023), UK Kidney Week (2023,2024), the International Immunosuppression & Transplant Skin Cancer Collaborative biennial symposium (2024), and the European Association of Dermato-Oncology congress (2024,2025).

## Data availability statement

The anonymised, compiled dataset supporting the findings of this study is available on request to MJB.

