## Supplemental Information for "Ongoing equipoise contributes to unchanged outcomes and significant morbidity following post-transplant cutaneous squamous cell carcinoma: A Multicentre Cohort Study"

**Supplementary Material**

**Table of Contents**

Supplementary Tables:

**Supplementary Table S1.** Full list of authors and affiliations

**Supplementary Table S2.** Data Collection Proforma

**Supplementary Table S3.** STROBE Checklist

**Supplementary Table S4.** Patients ineligible for analysis

**Supplementary Table S5.** Baseline characteristics of patients diagnosed with first cutaneous squamous cell carcinoma (CSCC), by contributing centre

**Supplementary Table S6**. Comparison of demographics with an international cohort

**Supplementary Table S7.** Comparison of first CSCC characteristics with an international cohort

**Supplementary Table S8.** Comparison of outcomes with an international cohort

Supplementary Figures

**Supplementary Figure S1.** Kaplan-Meir curves show event-free survival of international cohort

**Supplementary Tables**

| **Supplementary Table S1.** Full list of authors and affiliations | | |
| --- | --- | --- |
| **Name** | **Role** | **Affiliation** |
| Rachel Abbott | Consultant Dermatologist | Cardiff & Vale University Health Board, Cardiff, UK |
| Rabiah Ahmed | Specialty Trainee (Nephrology) | University Hospitals Birmingham NHS Foundation Trust, Birmingham, UK |
| Charles Archer | Consultant Dermatologist | Royal Berkshire NHS Foundation Trust, Reading, UK |
| Adarsh Babu | Consultant Nephrologist | Gloucestershire Hospitals NHS Foundation Trust, Gloucester, UK |
| Pippa Bailey | Consultant Nephrologist | North Bristol NHS Trust, Bristol, UK |
| Nitin Bhandary | Consultant Nephrologist | Royal Berkshire NHS Foundation Trust, Reading, UK |
| Matthew J. Bottomley* | Consultant Nephrologist | Oxford University Hospital NHS Foundation Trust, Oxford, UK |
| Jeva Cernova | Clinical Fellow (Dermatology) | Barts Health NHS Trust, London, UK |
| Tom Crisp | Internal Medicine Trainee | Oxford University Hospital NHS Foundation Trust, Oxford, UK |
| Amrit Darvay | Consultant Dermatologist | North Bristol NHS Trust, Bristol, UK |
| Jonathan Gamble | Specialty Trainee (Nephrology) | Cardiff & Vale University Health Board, Cardiff, UK |
| Maria Angela Gauci | Specialty Trainee (Nephrology) | North Bristol NHS Trust, Bristol, UK |
| Sian Griffin | Consultant Nephrologist | Cardiff & Vale University Health Board, Cardiff, UK |
| Catherine A Harwood | Consultant Dermatologist | Barts Health NHS Trust, London, UK |
| Emily Karn | Clinical Research Coordinator | Brigham & Women's Hospital, Boston, United States |
| Rubeta Matin | Consultant Dermatologist | Oxford University Hospital NHS Foundation Trust, Oxford, UK |
| James Moriarty | Consultant Nephrologist | Gloucestershire Hospitals NHS Foundation Trust, Gloucester, UK |
| Nina Muirhead | Associate Specialist (Dermatology) | Buckinghamshire Healthcare NHS Trust, Aylesbury, UK |
| Khizr Nawab | Specialty Trainee (Nephrology) | Royal Berkshire NHS Foundation Trust, Reading, UK |
| Alex Owen | Consultant Dermatologist | Gloucestershire Hospitals NHS Foundation Trust, Gloucester, UK |
| Emilia Peleva | Specialty Trainee (Dermatology) | Barts Health NHS Trust, London, UK |
| Emily Ruiz | Associate Professor (Dermatology) | Brigham & Women's Hospital, Boston, United States |
| Farida Shah | Consultant Dermatologist | University Hospitals Birmingham NHS Foundation Trust, Birmingham, UK |
| Adnan Sharif | Consultant Nephrologist | University Hospitals Birmingham NHS Foundation Trust, Birmingham, UK |
| Brenda Solomon | Clinical Research Assistant | Brigham & Women's Hospital, Boston, United States |
| Raj Thuraisingham | Consultant Nephrologist | Barts Health NHS Trust, London, UK |
| Harry Wakefield | Consultant Nephrologist | Royal Berkshire NHS Foundation Trust, Reading, UK |
| Elizabeth Wallin | Consultant Nephrologist | University Hospitals Birmingham NHS Foundation Trust, Birmingham, UK |
| Buddhika Wijayawickrama | Senior Clinical Fellow (Nephrology) | Barts Health NHS Trust, London, UK |

* Principal investigator

**Supplementary Table S2.** Data Collection Proforma

See separate file.

| Supplementary Table S3. STROBE Checklist | | |  |
| --- | --- | --- | --- |
|  | Item No | Recommendation | Page No |
| **Title and abstract** | 1 | (*a*) Indicate the study’s design with a commonly used term in the title or the abstract | 2-3 |
|  |  | (*b*) Provide in the abstract an informative and balanced summary of what was done and what was found | 3-4 |
| Introduction | | |  |
| Background/ rationale | 2 | Explain the scientific background and rationale for the investigation being reported | 5 |
| Objectives | 3 | State specific objectives, including any prespecified hypotheses | 6 |
| Methods | | |  |
| Study design | 4 | Present key elements of study design early in the paper | 6-7 |
| Setting | 5 | Describe the setting, locations, and relevant dates, including periods of recruitment, exposure, follow-up, and data collection | 7 |
| Participants | 6 | (*a*) Give the eligibility criteria, and the sources and methods of selection of participants. Describe methods of follow-up | 7 |
|  |  | (*b*) For matched studies, give matching criteria and number of exposed and unexposed | NA |
| Variables | 7 | Clearly define all outcomes, exposures, predictors, potential confounders, and effect modifiers. Give diagnostic criteria, if applicable | 7 |
| Data sources/ measurement | 8* | For each variable of interest, give sources of data and details of methods of assessment (measurement). Describe comparability of assessment methods if there is more than one group | 7 |
| Bias | 9 | Describe any efforts to address potential sources of bias | 7-9 |
| Study size | 10 | Explain how the study size was arrived at | 7 |
| Quantitative variables | 11 | Explain how quantitative variables were handled in the analyses. If applicable, describe which groupings were chosen and why | Supplementary table S2 |
| Statistical methods | 12 | (*a*) Describe all statistical methods, including those used to control for confounding | 8-9 |
|  |  | (*b*) Describe any methods used to examine subgroups and interactions | 9 |
|  |  | (*c*) Explain how missing data were addressed | Supplementary table S4 |
|  |  | (*d*) If applicable, explain how loss to follow-up was addressed | 7 |
|  |  | (*e*) Describe any sensitivity analyses | 8 |
| Results | | |  |
| Participants | 13* | (a) Report numbers of individuals at each stage of study—eg numbers potentially eligible, examined for eligibility, confirmed eligible, included in the study, completing follow-up, and analysed | 10 |
|  |  | (b) Give reasons for non-participation at each stage | Supplementary table S4 |
|  |  | (c) Consider use of a flow diagram | NA |
| Descriptive data | 14* | (a) Give characteristics of study participants (eg demographic, clinical, social) and information on exposures and potential confounders | 10-12  Table 2 |
|  |  | (b) Indicate number of participants with missing data for each variable of interest | The total numbers of recorded data for each variable are stated in each table |
|  |  | (c) Summarise follow-up time (eg, average and total amount) | 16 |
| Outcome data | 15* | Report numbers of outcome events or summary measures over time | 16-18  Table 4 |
| Main results | 16 | (*a*) Give unadjusted estimates and, if applicable, confounder-adjusted estimates and their precision (eg, 95% confidence interval). Make clear which confounders were adjusted for and why they were included | 16-19 |
|  |  | (*b*) Report category boundaries when continuous variables were categorized | NA |
|  |  | (*c*) If relevant, consider translating estimates of relative risk into absolute risk for a meaningful time period | NA |
| Other analyses | 17 | Report other analyses done—eg analyses of subgroups and interactions, and sensitivity analyses | 19-20 |
| Discussion | | | |
| Key results | 18 | Summarise key results with reference to study objectives | 20 |
| Limitations | 19 | Discuss limitations of the study, taking into account sources of potential bias or imprecision. Discuss both direction and magnitude of any potential bias | 24 |
| Interpretation | 20 | Give a cautious overall interpretation of results considering objectives, limitations, multiplicity of analyses, results from similar studies, and other relevant evidence | 21-23 |
| Generalisability | 21 | Discuss the generalisability (external validity) of the study results | 24 |
| Other information | | | |
| Funding | 22 | Give the source of funding and the role of the funders for the present study and, if applicable, for the original study on which the present article is based | 28 |

| **Supplementary Table S4.** Patients ineligible for analysis | |
| --- | --- |
| **Reasons for exclusion** | **No. of patients** |
| Care moved away from centre/abroad | 3 |
| First CSCC prior to study window | 8 |
| Non-cutaneous SCC | 2 |
| Patient receiving dialysis at time of CSCC | 2 |
| Incomplete data | 2 |
| CSCC, cutaneous squamous cell carcinoma; SCC, squamous cell carcinoma. | |

| **Supplementary Table S5.** Baseline characteristics of patients diagnosed with first cutaneous squamous cell carcinoma (CSCC), by contributing centre | | | | | | | | | |
| --- | --- | --- | --- | --- | --- | --- | --- | --- | --- |
| **Baseline Characteristics** | **All patients (n=136)** | **Centre A**  **(n=32)** | **Centre B**  **(n=12)** | **Centre C**  **(n=19)** | **Centre D**  **(n=8)** | **Centre E**  **(n=17)** | **Centre F**  **(n=23)** | **Centre G**  **(n=4)** | **Centre H**  **(n=21)** |
| Transplanting centre | NA | Yes | Yes | Yes | No | No | Yes | No | Yes |
| Age (years) | 64  (57-72) | 67  (55-71) | 63  (58-68) | 72  (58-74) | 60  (52-68) | 61  (57-70) | 65  (52-73) | 72  (69-73) | 62  (60-73) |
| Sex |  |  |  |  |  |  |  |  |  |
| Male | 102 (75.0) | 24 | 10 | 18 | 5 | 13 | 15 | 2 | 15 |
| Female | 34 (25.0) | 8 | 2 | 1 | 3 | 4 | 8 | 2 | 6 |
| Ethnicity |  |  |  |  |  |  |  |  |  |
| White | 128 (94.1) | 32 | 12 | 15 | 8 | 17 | 20 | 4 | 20 |
| Black | 2 (1.5) | 0 | 0 | 0 | 0 | 0 | 2 | 0 | 0 |
| Asian | 1 (0.7) | 0 | 0 | 0 | 0 | 0 | 0 | 0 | 1 |
| Not stated | 5 (3.7) | 0 | 0 | 4 | 0 | 0 | 1 | 0 | 0 |
| Previous SOM ^a^ | 11/135 | 6 | 1 | 1 | 0 | 1 | 0/22 | 1 | 1 |
| Previous skin cancer ^b^ | 46/135 | 14/31 | 3 | 4 | 2 | 6 | 11 | 3 | 3 |
| Previous actinic keratoses ^c^ | 67 (49.3) | 17 | 3 | 5 | 6 | 13 | 13 | 3 | 7 |
| Previous Bowen’s/keratoacanthoma ^d^ | 43/135 | 13 | 0 | 9 | 1 | 5 | 8 | 2 | 5 |
| Smoking history |  |  |  |  |  |  |  |  |  |
| Current | 11 (8.1) | 5 | 3 | 1 | 1 | 1 | 0 | 0 | 0 |
| Ex-smoker | 15 (11) | 7 | 3 | 1 | 0 | 1 | 1 | 2 | 0 |
| Never smoker | 37 (27.2) | 15 | 5 | 14 | 0 | 1 | 1 | 1 | 0 |
| Not recorded/unknown | 73 (53.7) | 5 | 1 | 3 | 7 | 14 | 21 | 1 | 21 |
| Total number of transplants |  |  |  |  |  |  |  |  |  |
| One | 107(78.7) | 27 | 11 | 16 | 7 | 11 | 17 | 3 | 15 |
| Two | 23 (16.9) | 4 | 1 | 2 | 0 | 4 | 6 | 1 | 5 |
| Three | 6 (4.4) | 1 | 0 | 1 | 1 | 2 | 0 | 0 | 1 |
| eGFR | 41  (27-55) | 33  (27-64) | 40  (29-58) | 44  (36-57) | 37  (26-55) | 42  (28-45) | 32  (23-44) | 38  (23-53) | 44  (33-60) |
| Previous rejection^e^ | 21/134 | 0/31 | 6 | 2 | 3 | 1 | 6 | 0 | 3/20 |
| Evidence of DSA prior to CSCC | 11/134 | 0 | 1 | 2 | 1 | 3 | 4 | 0 | 0 |
| Cumulative lifetime duration of IS (months) | 136  (68-204) | 166 (83-285) | 83  (62-100) | 149 (45-160) | 138  (48-185) | 120 (48-204) | 162 (115-216) | 83  (46-164) | 156 (93-252) |
| Induction therapies^e^ |  |  |  |  |  |  |  |  |  |
| Basiliximab/equivalent | 68/118 | 5/30 | 10 | 10/16 | 2/5 | 12 | 15/19 | 0 | 14/15 |
| Alemtuzumab | 21/118 | 10/30 | 0 | 1/16 | 3/5 | 5 | 0 | 2 | 0 |
| ATG/Thymoglobulin | 6/118 | 1/30 | 0 | 2/16 | 0 | 0 | 3/19 | 0 | 0 |
| Other | 4/118 | 0 | 1 | 3/16 | 0 | 0 | 0 | 0 | 0 |
| None | 19/118 | 14/30 | 1 | 0 | 0 | 0 | 1/19 | 2 | 1/15 |
| Number of IS agents at diagnosis^e^ |  |  |  |  |  |  |  |  |  |
| One | 8 (5.9) | 3 | 0 | 5 | 0 | 0 | 0 | 0 | 0 |
| Two | 70 (51.5) | 22 | 7 | 12 | 5 | 11 | 6 | 3 | 4 |
| Three | 58 (42.6) | 7 | 5 | 2 | 3 | 6 | 17 | 1 | 17 |
| Calcineurin/mTOR inhibitor |  |  |  |  |  |  |  |  |  |
| Tacrolimus | 104 (76.5) | 21 | 11 | 17 | 6 | 15 | 13 | 3 | 18 |
| Ciclosporin | 18 (13.2) | 6 | 0 | 0 | 1 | 1 | 8 | 0 | 2 |
| Sirolimus | 4 (2.9) | 1 | 0 | 0 | 1 | 0 | 1 | 1 | 0 |
| Nil | 10 (7.4) | 4 | 1 | 2 | 0 | 1 | 1 | 0 | 1 |
| Antiproliferative^e^ |  |  |  |  |  |  |  |  |  |
| MMF/MPA | 59 (43.4) | 10 | 1 | 10 | 3 | 7 | 15 | 3 | 10 |
| Azathioprine | 46 (33.8) | 17 | 5 | 2 | 4 | 1 | 5 | 1 | 11 |
| Nil | 31 (22.8) | 5 | 6 | 7 | 1 | 9 | 3 | 0 | 0 |
| Corticosteroid^e^ | 92 (67.6) | 13 | 12 | 6 | 4 | 16 | 21 | 1 | 19 |
| Year of diagnosis of first CSCC^e^ |  |  |  |  |  |  |  |  |  |
| 2016 | 29 (21.3) | 6 | 1 | 10 | 3 | 2 | 3 | 2 | 2 |
| 2017 | 20 (14.7) | 8 | 1 | 1 | 1 | 0 | 6 | 0 | 3 |
| 2018 | 27 (19.9) | 3 | 3 | 2 | 3 | 3 | 6 | 0 | 7 |
| 2019 | 36 (26.5) | 9 | 6 | 3 | 0 | 6 | 6 | 1 | 5 |
| 2020 | 24 (17.6) | 6 | 1 | 3 | 1 | 6 | 2 | 1 | 4 |
| Number of CSCC removed at first episode^e^ |  |  |  |  |  |  |  |  |  |
| Single | 127 (93.4) | 31 | 11 | 14 | 8 | 16 | 22 | 4 | 21 |
| Multiple | 9 (6.6) | 1 | 1 | 5 | 0 | 1 | 1 | 0 | 0 |
| Early management |  |  |  |  |  |  |  |  |  |
| IS reduction | 39 (28.7) | 9 | 3 | 3 | 4 | 1 | 8 | 1 | 10 |
| Topical chemoprevention | 28 (20.6) | 0 | 7 | 5 | 0 | 6 | 9 | 0 | 1 |
| Systemic chemoprevention | 6 (4.4) | 0 | 3 | 0 | 0 | 3 | 0 | 0 | 0 |
| Values are expressed as number (percentage) or median (interquartile range), as appropriate.  Where data is missing, results have been expressed as a fraction.  ATG, anti-thymocyte globulin; CSCC, cutaneous squamous cell carcinoma; DSA, donor specific antibody; eGFR, estimated glomerular filtration rate; IS, immunosuppression; MMF, mycophenolate mofetil; MPA, mycophenolic acid; mTOR, mammalian target of rapamycin, SOM: solid organ malignancies.  ^a^10/11 cancers were in remission at study baseline.  ^b^ Previous basal cell carcinoma (n=44) and/or melanoma(n=3) .  ^c^ Includes both clinical and histological diagnoses.  ^d^ Previous Bowen’s disease (CSCC-in situ, histologically diagnosed in 37 patients and clinically diagnosed by a dermatologist in 4 patients) and/or keratoacanthoma (histologically diagnosed in 3 patients)  ^e^ Significant differences between centres (*P* <0.05), based on chi-squared test. | | | | | | | | | |

| **Supplementary Table S6**. Comparison of demographics with an international cohort | | |
| --- | --- | --- |
|  | **COAST** | **International** |
| **Baseline Characteristics** | **All patients (n=136)** | **All patients (n=79)** |
| Age (years), median (IQR) | 64 (57-72) | 62 (54-70) |
| Sex |  |  |
| Male | 102 (75.0) | 53 (67.1) |
| Female | 34 (25.0) | 26 (32.9) |
| Race |  |  |
| White | 128/131 (97.7) | 78/78 (100) |
| Black | 2/131 (1.5) | - |
| Asian | 1/131 (0.7) | - |
| Previous skin cancer | 46/135 (34.1) | 31 (39.2) |
| Total number of transplants |  |  |
| One | 107 (78.7) | 74 (93.7) |
| Two | 23 (16.9) | 5 (6.3) |
| Three | 6 (4.4) | - |
| Types of Transplant |  |  |
| Kidney | 136 (100) | 46 (57.5) |
| Lung | - | 22 (27.5) |
| Heart | - | 11 (13.7) |
| Liver | - | 3 (3.8) |
| Pancreas | - | 2 (2.5) |
| eGFR^a^ | 41 (27-55) | 32 (21-56) |
| Cumulative lifetime duration of IS (months) | 136 (68-204) | 63.5 (31.7-162.0) |
| Number of IS agents at diagnosis |  |  |
| One | 8 (5.9) | 1 (1.3) |
| Two | 70 (51.5) | 32 (40.0) |
| Three | 58 (42.6) | 46 (57.5) |
| Calcineurin/mTOR inhibitor |  |  |
| Tacrolimus | 104 (76.5) | 62 (78.5) |
| Cyclosporin | 18 (13.2) | 8 (10.1) |
| Sirolimus | 4 (2.9) | 4 (5.1) |
| Everolimus | 0 | 1(1.2) |
| Nil | 10 (7.4) | 4 (5.1) |
| Antiproliferative |  |  |
| MMF/MPA | 59 (43.4) | 39 (48.4) |
| Azathioprine | 46 (33.8) | 26 (32.9) |
| Nil | 31 (22.8) | 14 (17.7) |
| Corticosteroid  Prednisone | 92 (67.6) | 60 (75.9) |
| Number of CSCC removed at first episode |  |  |
| Single | 127 (93.4) | 57/69 (82.6) |
| Multiple | 9 (6.6) | 12/69 (17.4) |
| Values are expressed as number (percentage), or median (interquartile range), as appropriate.  CSCC, cutaneous squamous cell carcinoma; DSA, donor specific antibody; eGFR, estimated glomerular filtration rate; IS, immunosuppression; MMF, mycophenolate mofetil; MPA, mycophenolic acid; mTOR, mammalian target of rapamycin.  ^a^ only collected for patients with kidney transplants | | |

| **Supplementary Table S7.** Comparison of first CSCC characteristics with an international cohort | | |
| --- | --- | --- |
|  | **COAST** | **International** |
| **Characteristics** | **All tumours (n=147)** | **All tumours (n=95)** |
| Location |  |  |
| Head and neck | 93 (63.3) | 52/94 (55.3) |
| Trunk | 12 (8.2) | 11/94 (11.7) |
| Upper limb | 37 (25.2) | 19/94 (20.2) |
| Lower limb | 5 (3.4) | 12/94 (12.8) |
| Diameter (mm) | 12 (9-20) | 11 (6.0-15.0) |
| Depth (mm) | 3 (1.9-5) | 15 (8.0-30.0) |
| Invasion beyond the subcutis | 8/128 (6.3) | 8 (8.4) |
| Differentiation |  |  |
| Well | 40/145 (27.6) | 72/85 (84.7) |
| Moderate | 78/145 (53.8) | 10/85 (11.8) |
| Poor | 27/145 (18.6) | 3/85 (3.5) |
| PNI | 8/130 (6.2) | 6 (6.3) |
| LVI | 3/141 (2.1) | 2 (2.1) |
| Clear margins after initial excision | 117/146 (80.1) | 84 (88.5) |
| BWH |  |  |
| T1 | 77/133 (57.9) | 67/85 (78.8) |
| T2a | 41/133 (30.8) | 10/85 (11.8) |
| T2b | 15/133 (11.3) | 6/85 (7.1) |
| T3 | - | 2/85 (2.4) |
| AJCC8/UICC8 |  |  |
| T1 | 91/134 (67.9) | 68/85 (80.0) |
| T2 | 11/134 (8.2) | 11/85 (12.9) |
| T3 | 32/134 (23.9) | 6/85 (7.1) |
| Values are expressed as number (percentage) or median (interquartile range), as appropriate.  AD-SCI, actinic damage and skin cancer index; AJCC8/UICC8, the 8th edition of the American Joint Committee on Cancer/International Union Against Cancer staging systems; BWH, Brigham and Women’s Hospital classification; LVI, lympho-vascular invasion; PNI, perineural invasion. | | |

| **Supplementary Table S8.** Comparison of outcomes with an international cohort | | | |
| --- | --- | --- | --- |
|  | **COAST** | | **International** |
| **Outcome** | | **All patients (n=136)** | **All patients (n=79)** |
| Follow up (months) | | 39 (26-52) | 59 (38.0-93.0) |
| Poor outcomes ^a^ | | 99 (72.8) | 40 (50.6) |
| Death | | 38 (23.3) | 32 (40.5) |
| Time to death (months) | | 20 (12-32) | 51.0 (35.3-65.0) |
| Causes of death | |  |  |
| CSCC-related | | 11 (28.9) | 3 (9.4) |
| CVD | | 6 (15.8) | 2 (6.2) |
| ARDS | | - | 5 (15.6) |
| Infection | | 6 (15.8) | 1 (3.2) |
| Other malignancy | | 2 (5.3) | 3 (9.3) |
| Other | | 2 (5.3) | - |
| Unknown | | 10 (26.3) | 17 (53.1) |
| Graft loss | | 17 (12.5) | 7 (8.8) |
| Time to graft loss (months) | | 34 (18-45) | 28.0 (22.0-31.0) |
| Further CSCC | | 66 (48.5) | 60 (75.9) |
| Median time to further CSCC (months) | | 13 (8-27) | 18.0 (8.7-46.5) |
| Metastatic CSCC | | 18/135 (13.3) | 10 (12.6) |
| Time to metastatic CSCC (months) | | 11 (7-12) | 29.0 (6.5-62.5) |
| Solid-organ malignancy | | 10 (7.4) | 4 (5.1) |
| Time to solid-organ malignancy (months) | | 16 (14-27) | 53 (35.5-68.5) |
| Values are expressed as number (percentage) or median (interquartile range), as appropriate.  ARDS, Acute respiratory distress syndrome; CSCC, cutaneous squamous cell carcinoma; CVD, cardiovascular disease.  ^a^ Composite outcome including graft loss, further CSCC, solid-organ malignancy, metastatic CSCC, or death with a functioning graft. | | | |

**Supplementary Figures**

**Supplementary Figure S1.** Kaplan-Meir curves show event-free survival of international cohort

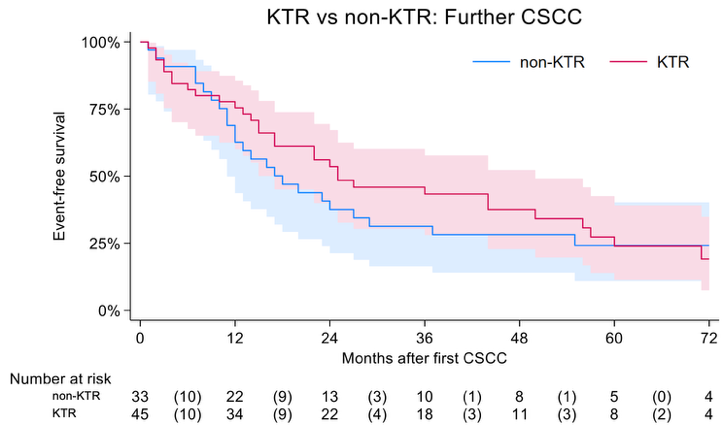

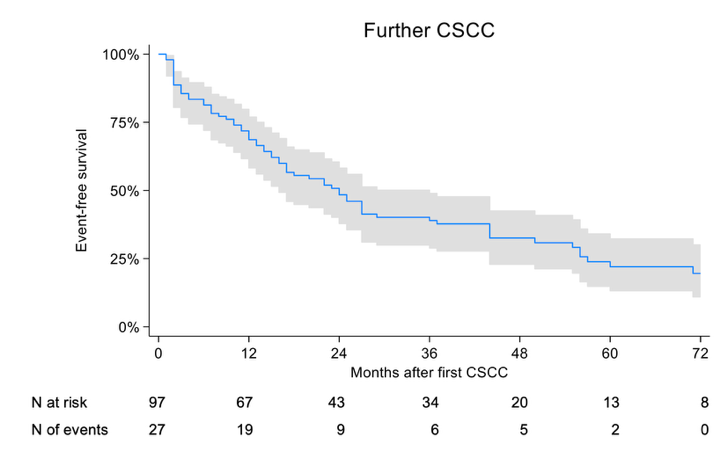

b

a

d

c

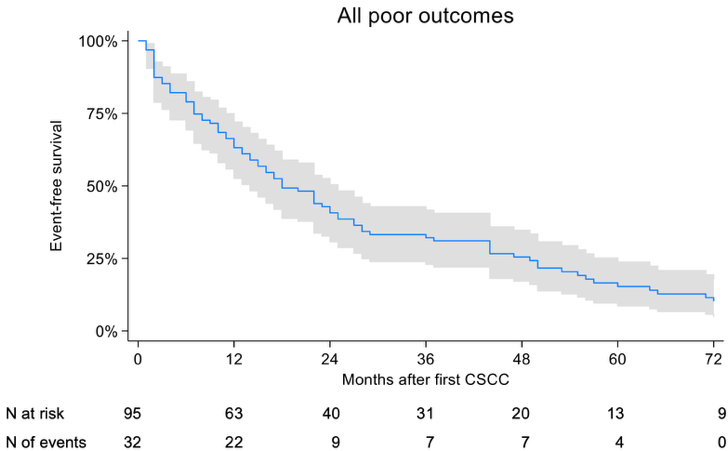

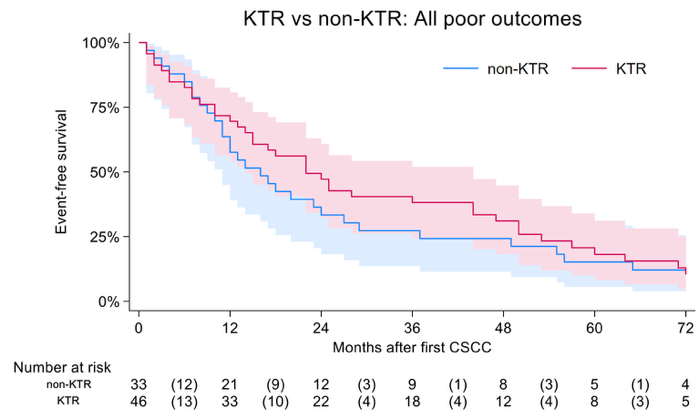

e

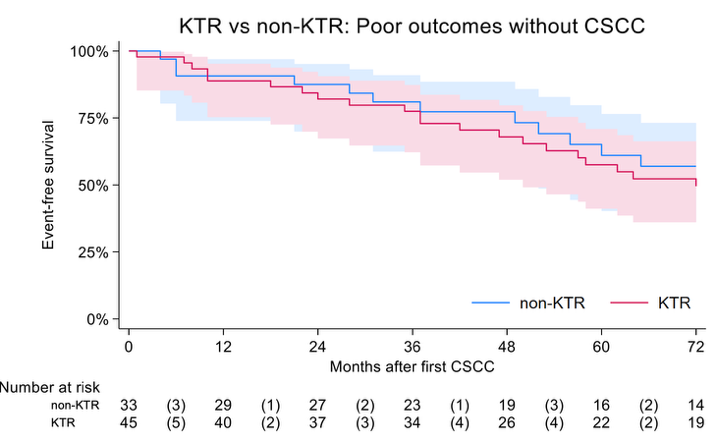

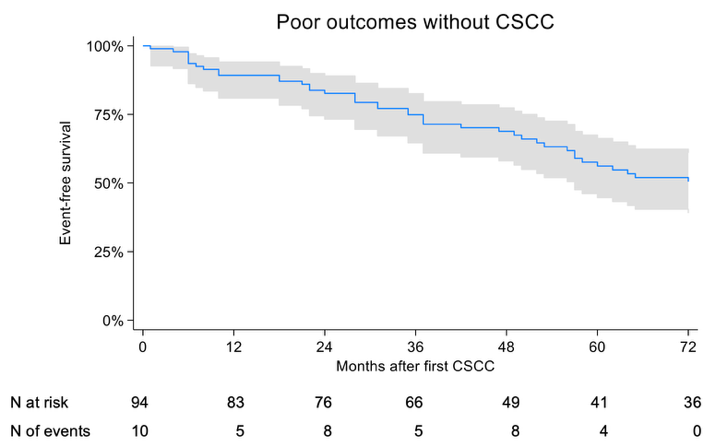

f

Kaplan-Meier curves show time to further CSCC for (a) the entire cohort and (b) stratified by transplant type: kidney transplant recipients (KTR) versus recipients of other organs (non-KTR). Kaplan-Meier curves show time to the composite ‘all poor outcomes’ for (c) the entire cohort and (d) stratified by transplant type. Kaplan-Meier curves show time to ‘poor outcomes without CSCC’ for (e) the entire cohort and (f) stratified by transplant type. CSCC: cutaneous squamous cell carcinoma; KTR: kidney transplant recipient.
