## Supplementary material for "Ongoing equipoise contributes to unchanged outcomes and significant morbidity following post-transplant cutaneous squamous cell carcinoma: A Multicentre Cohort Study": Figure Legends

**Figure 1 Baseline characteristics and early management after a first cutaneous squamous cell carcinoma (CSCC).**(a) Pie chart shows number of patients per centre, with centres labelled A-H; 136 patients were included in total. Stacked bar plots show baseline characteristics at time of first CSCC, including (b) number of immunosuppressive agents, (c) antiproliferative agents used, and (d) BWH stage of first CSCC. Results are shown for the entire cohort (All) and for each centre (A-H). (e) Stacked bar plot shows immunosuppression reduction approach for the entire cohort (‘All’) and by centre. Overall, immunosuppression was reduced in 28.7% of patients (range 5.9-50.0% by centre). (f) Bar plot shows initiation of topical therapy for the entire cohort (20.6%) and by centre (range 0-58.3%). 5-fluorouracil cream was started in all but one case, in whom imiquimod cream was used. BWH, Brigham and Women’s Hospital classification; CNI, calcineurin inhibitor; CSCC, cutaneous squamous cell carcinoma; MMF, mycophenolate mofetil; MPA, mycophenolic acid.

**Figure 2 Outcomes after first cutaneous squamous cell carcinoma (CSCC) in kidney transplant recipients (KTR).** (a) Alluvial plot shows sequence of events after the first CSCC, including graft loss, death with a functioning graft (DWFG), metastasis, new solid-organ malignancy (SOM), further CSCC, or censoring due loss to follow-up (LTFU) or study end. (b) Stacked ridgeline plot shows time from diagnosis of first CSCC to development of poor outcomes and to last follow-up (last clinic appointment before loss to follow-up or study end). (c) Kaplan-Meier curve shows time to further CSCC. (d) Bar plot shows cumulative incidence of further CSCC at 1, 3 and 5 years after a first-ever CSCC in KTR in the present study (COAST) and in historical published cohorts. The dashed lines show the weighted averages, based on the number of participants in each study. (e)Kaplan-Meier curve shows time to the composite ‘all poor outcomes’. (e)Kaplan-Meier curve shows time to ‘poor outcomes without CSCC’. CSCC, cutaneous squamous cell carcinoma; DWFG, death with a functioning graft; KTR, kidney transplant recipients; LTFU, loss to follow-up; SOM, solid organ malignancy.

**Figure 3 Univariate and multivariate modelling results.** Forest plots showing results for risk of further CSCC from (a) univariate modelling and from multivariate modelling analysis with competing risk analysis using the Fine-Gray method, including either (b) BWH or (c) AD-SCI grading (multiple first CSCC is already part of AD-SCI and was not included as a separate variable). Forest plots show results for risk of any poor outcome from (d) univariate modelling and multivariate modelling analysis, including either (e) BWH or (f) AD-SCI. Forest plots show results for risk of any ‘poor outcome without CSCC’ from (g) univariate modelling and (h) multivariate modelling analysis including BWH. Univariate modelling results are shown for all significant variables, as well as for important non-significant variables. Variables that were statistically significant or showed a trend towards significance (P <0.10) on univariate analyses, as well as pre-selected clinically-important risk factors (age and/or duration of immunosuppression) were included in the multivariate models, whilst smoking status and history of pre-malignant skin disease were excluded due to underreporting. Variables included baseline characteristics: Male sex, Age at first CSCC, Duration of immunosuppression (in months), Smoker (current vs never), Previous solid-organ malignancy, Previous BCC, AK/Bowen’s/KA (history of premalignant skin lesions), Multiple first CSCC, BWH (grades≥T2b), AJCC8/UICC8 (high-risk first CSCC, defined as grades T3 or greater), AD-SCI (grades 5 or 6), and BAD (‘Very high’ risk). Significant results (P < 0.05) are indicated with an asterisk. AD-SCI, Actinic Damage and Skin Cancer Index; AJCC8/UICC8, 8th edition of the American Joint Committee on Cancer/International Union Against Cancer staging system; AK, actinic keratosis; BAD, British Association of Dermatologists’ SCC guidelines; BWH, Brigham and Women’s Hospital classification; CSCC, cutaneous squamous cell carcinoma; CI, confidence interval; KA, keratoacanthoma; HR, hazard ratio; SOM, solid organ malignancy.
